# Comparing Interventions Aimed at Improving Influenza Vaccination Coverage Among Vulnerable Populations: A Systematic Review and Meta-Analysis Protocol

**DOI:** 10.1101/2021.03.08.21253172

**Authors:** Pei Chen Wu, Lyn McPherson, Stephen B Lambert, Peter Wnukowski-Mtonga, Nicholas G Lennox, Robert S Ware

## Abstract

**Background:** Influenza is a major contributor to global disease burden. Vaccination recommendations specifically target populations at increased risk of serious influenza sequelae. The aim of this study is to conduct a systematic review and meta-analysis to evaluate the effectiveness of different quality improvement interventions to increase vaccination rates in high-risk populations.

**Methods:** Randomized and nonrandomized studies with concurrent control groups will be identified. Interventions to increase vaccination rates will be categorized by strategy type. Overall intervention effects will be calculated using random effects models. Study quality will be assessed using a modified Cochrane Risk of Bias tool.

## BACKGROUND

Influenza is a vaccine-preventable acute respiratory infection directly responsible for three to five million cases of severe illness annually, resulting in 250,000 to 500,000 deaths.^1^ The financial burden of influenza and its sequelae are substantial: estimates from the United States are of annual direct losses of US$10.4 billion as a direct consequence of influenza, with an additional US$16.3 billion lost in sick days and productivity.^2^ Influenza disproportionately affects infants, young children, adults aged 65 years or older, pregnant women, immunocompromised individuals, First Nations peoples, and those suffering from chronic disease; all of whom are at risk of experiencing increased morbidity and mortality due to influenza.^3,4^ Current health care guidelines in most developed nations recommend annual vaccination for the aforementioned high-risk groups. In order to maximize the benefits these populations derive from immunization, the US Department of Health and Human Services has launched the Healthy People 2020 initiative in order to improve preventive health care.^5^ The initiative aims to increase influenza vaccination rates for various high-risk groups to targets ranging from 70 to 90 percent. Vaccination coverage rates for vulnerable populations have been rising substantially over the past two decades in most developed countries. In Australia, between 1 March and 19 April 2020, over 2.1 million influenza vaccinations were administered and recorded in the Australian Immunisation Register compared to 624,000 at the same time in 2019 and 235,000 in 2018^6^, as government, GPs and other vaccine providers encouraged people to get vaccinated to avoid the combination of COVID-19 and influenza which could be life threatening, but inconsistent and even decreasing trends in coverage have been recently reported in the U.S,^7^ Europe^8^ and Canada,^9^ indicating that coverage rates still fall short of the goals proposed by the Healthy People 2020 initiative.

In developed nations, the majority of influenza vaccinations take place in community settings. community-based primary care centers remain the most effective way to promote health to the public, and various outreach initiatives have been trialled in the community with the aim to increase influenza vaccination coverage in vulnerable populations. Ompad et al.^10^ previously reviewed over 50 studies examining interventions directed towards high-risk populations, and found the majority of interventions did not result in vaccination rates that met the Healthy People 2010 goals. Moreover, the study did not perform statistical analyses to evaluate the effectiveness of each intervention, and did not focus on community-settings. A previous systematic review and meta-analysis by Lau et al.^11^ found that various quality improvement interventions were effective in increasing influenza and pneumococcal vaccination coverage in elderly adults, but did not examine intervention effects on other vulnerable populations, and did not focus exclusively on primary care interventions.

The aim of this study is to systematically evaluate, compare, and analyze the effectiveness of different intervention strategies for improving influenza vaccination coverage, with the goal of identifying the most effective strategies to improve vaccination coverage among vulnerable populations in the community.

This review protocol will follow the guidelines for the Preferred Reporting Items for Systematic Reviews and Metanalysis (PRISMA)^12^ and is reported here using the Guidance notes for registering a systematic review protocol with PROSPERO provided by the Centre for Reviews and Dissemination.^13^

## PROSPERO ITEMS

### 1. Review title

Comparing Interventions Aimed at Improving Influenza Vaccination Coverage Among Vulnerable Populations: A Systematic Review and Meta-Analysis

### 2. Original language title

As above

### 3. Anticipated or actual start date

15 March 2021

### 4. Anticipated completion date

30 September 2021

### 5. Stage of review at time of this submission

This review topic was first explored in December 2016/January 2017 and then again in December 2017/January 2018 as part of a University of Queensland Summer Scholarship Program. During this time the search strategy was developed and the publications appearing prior to December 2017 were identified, the study characteristics were recorded, and data was extracted. After this Protocol is submitted the search will be re-run and eligible publications will be identified for inclusion in meta-analyses.

### 6. Named contact

Prof Robert Ware

### 7. Named contact email

; https://experts.griffith.edu.au/18978-robert-ware

### 8. Named contact address

Centre for Applied Health Economics, Griffith University, 170 Kessels Road, Nathan QLD 4111, AUSTRALIA

### 9. Named contact phone number

+61 7 3735 9117

### 10. Organisational affiliation

Menzies Health Institute Queensland, Griffith University

### 11. Review team members and their organisation affiliations

Pei Chen Wu; Faculty of Medicine, The University of Queensland; Queensland Centre for Intellectual and Developmental Disability, The University of Queensland

Lyn McPherson; Menzies Health Institute Queensland, Griffith University; Queensland Centre for Intellectual and Developmental Disability, The University of Queensland

Stephen B Lambert, Faculty of Medicine, The University of Queensland

Peter Wnukowski-Mtonga; National Centre for Immunisation Research & Surveillance, The Children’s Hospital at Westmead

Nicholas G Lennox; Queensland Centre for Intellectual and Developmental Disability, MRI-UQ, The University of Queensland

Robert S. Ware; Menzies Health Institute Queensland, Griffith University; Queensland Centre for Intellectual and Developmental Disability, The University of Queensland

## Data Availability

This is a protocol for a systematic review. Data will be extracted from identifed articles. Extracted data will be reported in published article

## 14. Collaborators

Nil

## 15. Review question

Which vaccination strategies are the most effective for improving influenza vaccination coverage among vulnerable populations in the community?

## 16. Searches

We will identify relevant studies for synthesis using four databases (Cochrane Library via John Wiley & Sons Inc., MEDLINE via EBSCOHost, EMBASE via Elsevier, and CINAHL via EBSCOHost). Search strategies for each database were prepared by PW in consultation with other team members and a university library Information specialist. Preliminary searches were conducted using the various suggested terms to pilot the study selection process. After screening search results against eligibility criteria, modifications were made as considered necessary.

## 17. URL to search strategy

The final search strategies as used in the first version of this systematic review are detailed in Appendix I.

## 18. Condition or domain being studied

Influenza vaccination

## 19. Participants/population

Studies will be included if they compared the effectiveness of a quality improvement intervention against a contemporaneous comparison group in a community setting. For inclusion, studies will need to report vaccination rates, be published in an English language peer-reviewed journal and involve a high-risk population living in the community. A population is defined as being high risk if it was included in the guidelines provided by the relevant health authorities in the USA, Australia, Canada, or the United Kingdom (See Appendix 2).

## 20. Interventions/exposures

Strategies will be categorized based on the classification system proposed by Shojania et al.^14^ and subsequently adapted by Lau et al.^11^. Categories will be audit and feedback, case management, clinician education, clinician reminders, community engagement, continuous quality improvement, delivery site change, financial incentives for clinicians, financial incentives for patients, patient outreach, changes in care team personnel (team change), and changes to patient visit routine (visit structure change). Where appropriate, quality improvement strategies with a sufficient number of comparisons will be organized into sub-categories, and meta-analyses will be conducted within each sub-category.

## 21. Comparator/control

Comparator group will be either usual care, non-vaccination intervention, or, when intervention group received additional strategy, whatever strategy the control group received.

## 22. Types of study to be included

Randomised Controlled Trials (RCTs) and quasi RCTs,

## 23. Context

Studies need to be conducted among groups living in the community.

## 24. Main outcomes

Vaccination rate

## 25. Additional outcomes

Nil

## 26. Data extraction (selection and coding)

The bibliographic software, EndNote, will be employed to organize, store, and manage all the citations. References retrieved from the four databases will be imported into Endnote. After removal of duplicate references, reviewers (PW and RW) will select studies for inclusion in the meta-analyses, first by screening of the titles and abstracts, and secondly, using full texts. We will attempt to obtain any references not available on-line or locally with the assistance of the University Library. Any differences in the selection of studies for inclusion will be resolved by discussion between reviewers and the other members of the research team if necessary. PW will extract data from each selected study.

## 27. Risk of bias (quality) assessment

Study quality will be measured using a modified version of the Cochrane Risk of Bias (ROB) tool,^15^ a 6-item questionnaire assessing the appropriateness of random sequence generation, allocation concealment, blinding of participants, personnel, and outcome assessors, data completeness, and other sources of biases. For quality assessment we summed individual scores.

## 28. Strategy for data synthesis

All studies reporting sufficient data to derive odds ratios and standard errors will be included in meta-analyses. Comparisons will be eligible to be included in the meta-analysis if the control group received usual care, or if the control intervention was focused towards non-vaccination behaviour, or if the intervention group received an additional strategy as well as whatever strategy was applied to both the intervention and control groups. When a given intervention can be classified into multiple quality improvement strategies, the number of subjects in the intervention group will divided between the strategies to avoid double-counting participants. Results from individual studies will be synthesized using random-effects models. Results will be reported overall, and by quality improvement strategy. Heterogeneity will be measured using the I^2^ statistic. The number needed to target for vaccination will be calculated. Sensitivity analyses will be conducted to investigate any association between study quality (assessed by ROB scores) and reported effect sizes using meta-regression. Publication bias will be tested by generating funnel plots and applying Harbord’s test.^16^ Analyses will be performed using Stata v14.0 (StataCorp LP, College Station, TX, USA).

## 29. Analysis of subgroups or subsets

Nil

## 30. Type and method of review

Type of review - Systematic review and meta-analysis

Health area of review – Public Health

## 31. Language

English

## 32. Country

Australia

## 33. Other registration details

Nil

## 34. References

See below

## 35. Dissemination plans

It is intended to publish the results of this study in a peer-reviewed journal. Any amendments to this proposal will be documented and communicated in the final published manuscript.

## 37. Details of any existing review of the same topic by the same authors

**Nil**

## 38. Current review status

Ongoing

## 39. Any additional information

Nil

## Appendix I: Database Search Strategies

### MedLine

**Search performed on November 18th, 2016; returning 1090 articles**.

**# Query**

1. (MH “Influenza Vaccines”)
2. MH vaccines
3. MH immunization
4. vaccin*
5. “immuni*”
6. “inoculat*”
7. “prevent*”
8. 1 OR 2 OR 3 OR 4 OR 5 OR 6 OR 7
9. (MH “Influenza, Human”)
10. (MH “Influenza B virus”) OR (MH “Influenza A virus”) OR (MH “Influenza C virus”)
11. (MH “Influenzavirus A”) OR (MH “Influenzavirus B”) OR (MH “Influenzavirus C”)
12. AB influenza OR TI influenza
13. AB flu OR TI flu
14. 9 OR 10 OR 11 OR 12 OR 13
15. AB primary care OR TI primary care
16. AB General practi* OR TI General practi*
17. AB Primary health* OR TI Primary health*
18. AB Community mental health* OR TI Community mental health*
19. AB Family practice OR TI Family practice
20. AB Family medicine OR TI Family medicine
21. AB Family physician* OR TI Family physician*
22. TI Home care OR AB Home care
23. AB Home based OR TI Home based
24. AB Home health* OR TI Home health*
25. AB Community health* OR TI Community health*
26. AB Community nurs* OR TI Community nurs*
27. AB health visit* OR TI health visit*
28. AB Community pharmac* OR TI Community pharmac*
29. AB Preventive care OR TI Preventive care
30. AB Prevention program* OR TI Prevention program*
31. AB Preventive OR TI Preventive
32. 15 OR 16 OR 17 OR 18 OR 19 OR 20 OR 21 OR 22 OR 23 OR 24 OR 25 OR 26 OR 27 OR 28 OR 29 OR 30 OR 31
33. PT randomized controlled trial
34. “random*”
35. “control*”
36. “intervention*”
37. “evaluat*”
38. “compar*”
39. “impact”
40. 33 OR 34 OR 35 OR 36 OR 37 OR 38 OR 39
41. “rates”
42. “rate”
43. “distribution”
44. “coverage”
45. “status”
46. “delivery”
47. “program”
48. “intervention”
49. “promotion”
50. “initiative”
51. “strategy”
52. “uptake”
53. 41 OR 42 OR 43 OR 44 OR 45 OR 46 OR 47 OR 48 OR 49 OR 50 OR 51 OR 52
54. MH humans
55. 8 AND 14 AND 32 AND 40 AND 53 AND 54

Limiters - English Language; Age Related: Infant, Newborn: birth-1 month, Infant: 1-23 months, All Infant: birth-23 months, Child, Preschool: 2-5 years, Young Adult: 19-24 years, Adult: 19-44 years, Middle Aged: 45-64 years, Middle Aged + Aged: 45 + years, Aged: 65+ years, Aged, 80 and over, All Adult: 19+ years

**Update: Search performed on Nov 27th, 2017; returning 1**,**189 results. Filter for papers published between 2016-2017 yielded an additional 109 results**.

### Embase

**Search performed on November 18th, 2016; returning 1149 articles.**

**# Query**

1. (‘influenza’ OR influenza OR flu) AND ‘influenza’ OR influenza OR flu
2. ‘influenza vaccines’/exp OR ‘vaccine’/exp OR ‘immunization’/exp OR vaccin* OR immuni* OR inoculat* OR prevent*
3. rates OR rate OR uptake OR determinant* OR tool* OR distribution OR coverage OR status OR delivery OR program* OR interven* OR promot* OR initiat* OR strateg*
4. ‘primary care’/exp OR ‘primary health care’/exp OR ‘primary health*’:ab,ti OR ‘general practise’ OR ‘general practi*’:ab,ti OR ‘family practice’:ab,ti OR ‘family physician*’ OR ‘family medicine’:ab,ti OR ‘community mental health’:ab,ti OR ‘community mental health services’:ab,ti OR ‘home care services’/exp OR ‘home care’:ab,ti OR ‘home based’:ab,ti OR ‘home health*’:ab,ti OR ‘health visit*’:ab,ti OR ‘health workers’ OR ‘community nurs*’:ab,ti OR ‘community pharmac*’:ab,ti OR ‘community health*’:ab,ti OR ‘community health services’/exp OR ‘preventive health services’/exp OR ‘preventive care’:ab,ti OR ‘prevention program*’:ab,ti
5. randomized:it AND controlled:it AND trial:it OR random* OR control* OR intervention* OR compar* OR impact OR evaluat*
6. ‘influenza’/exp OR ‘influenza b virus’/exp OR ‘influenzavirus a’/exp OR ‘influenzavirus c’/exp OR influenza:ab,ti OR flu:ab,ti
7. [english]/lim AND [humans]/lim
8. #1 AND #2 AND #3 AND #4 AND #5 AND #6 AND #7
9. [adult]/lim OR [aged]/lim OR [infant]/lim OR [middle aged]/lim OR [newborn]/lim OR [preschool]/lim OR [very elderly]/lim OR [young adult]/lim
10. #8 AND #9

**Update: Search performed on Nov 27th, 2017; returning 1**,**215 results. Filter for papers published between 2016-2017 yielded an additional 136 results**.

### Cochrane

**Search performed on November 18th, 2016; returning 1283 articles**.

**# Query**

1. (immuniz* or immunis* or inoculat* or vaccin*):ti,ab,kw and (influenza):ti,ab,kw (Word variations have been searched)
2. rate or rates or uptake or delivery or distribution or coverage or status:ti,ab,kw or “quality improvement” or qi or “quality management” or “case management” or “patient care” or interdisciplinary:ti,ab,kw or registr* or audit or education or reminder* or “self care” or “self management” or “medical record” or program* or intervention*:ti,ab,kw or “primary care”:ti,ab,kw (Word variations have been searched)
3. #1 and #2

**Update: Search performed on Nov 27th, 2017; returning 1**,**453 results. Filter for papers published between 2016-2017 yielded an additional 216 results**.

### CINAHL

**Search performed on November 18th, 2016; returning 140 articles**.

**# Query**

1. MH immunization
2. TI vaccin* OR AB vaccin*
3. TI immunis* OR AB immunis*
4. TI immuniz* OR AB immuniz*
5. TI inoculat* OR AB inoculat*
6. MH Influenza
7. TI influenza OR AB influenza
8. MH Influenza Vaccine
9. S1 OR S2 OR S3 OR S4 OR S5
10. S6 OR S7
11. S9 AND S10
12. S8 OR S11
13. S8 OR S11

Limiters - English Language; Exclude MEDLINE records; Human; Age Groups: Infant, Newborn: birth-1 month, Infant: 1-23 months, Child, Preschool: 2-5 years, Adult: 19-44 years, Middle Aged: 45-64 years, Aged: 65+ years, Aged, 80 and over, All Infant, All Adult

**Update: Search performed on Nov 27th, 2017; returning 209 results. Filter for papers published between 2016-2017 yielded an additional 92 results**.

